# Significance of Right Ventricular Myocardial Work for Clinical Improvement in Heart Failure with Reduced Ejection Fraction Patients after Transcatheter Edge-To-Edge Repair

**DOI:** 10.1101/2024.03.01.24303636

**Authors:** Jie Zeng, Qinglan Shu, Yi Liu, Cong Lu, Yun Xu, Yi Zhou, Qingfeng Zhang, Luwei Ye, Qingguo Meng, Sijia Wang, Yuan Yao, Xinyi Lin, Yi Wang

## Abstract

**Aims:** It remains uncertain which patients would benefit the most from transcatheter edge-to-edge repair (TEER). We aim to investigate the relationship between right ventricular function, as assessed by pressure-strain loops (PSL), and post-TEER clinical improvement.

**Methods and results:** A total of 48 heart failure with reduced ejection fraction (HFrEF) patients (68±15 yrs) with moderate-to-severe or severe SMR were enrolled for TEER. Impaired health status (Kansas City Cardiomyopathy Questionnaire Overall Summary Score [KCCQ-OS]) and exercise capacity (6-min walk distance [6MWD]) were evaluated at baseline and during 1-year follow up. Before and right after TEER, myocardial work (MW) metrics were non-invasively evaluated, including global work index (GWI), global constructive work (GCW), global wasted work (GWW), and global work efficiency (GWE). RV GLS, RVGWI, RVGCW, RVGWE were significantly increased after MitraClip treatment (-9.7±3.8%, 452.4±112.5 mmHg%, 596.3±127.5 mmHg% and 85.7±15.6% before vs -12.5±3.5%, 589.4±119.6 mmHg%, 778.8±135.3 mmHg% and 91.2±22.4% after MitraClip treatment, *p* = 0.025, 0.030, 0.025 and 0.037, respectively). The Kaplan-Meier estimates for survival, freedom from HF hospitalization at 12 months were 95.8% and 89.1%. On multivariable linear regression analysis, RVGWI and RVGCW immediate change was independently associated with KCCQ-OS (△RVGWI: ***β*** = 0.40, *P* < 0.001; △RVGCW: ***β*** = 0.39, *P* =0.003), RVGWI, RVGCW and RVGLS immediate change were independently associated with 6MWD improvement (△RVGWI: ***β*** = 0.31, *P* = 0.029; △RVGCW: ***β*** = 0.30, *P* = 0.039; △RVGLS: ***β*** = 0.35, *P* = 0.041).

**Conclusion:** RVMW was significantly increased after MitraClip treatment. And RV reserve function is an important predictor of clinical improvement in HFrEF patients with TEER.

## Introduction

Heart failure (HF), a leading cause of cardiac-related mortality, continues to present significant challenges worldwide. A substantial proportion of these patients have heart failure with reduced ejection fraction (HFrEF) and concomitant significant secondary mitral regurgitation (SMR), contributing to worsening symptoms and poor prognosis. The COAPT Clinical Trials have demonstrated promising results through the use of transcatheter edge-to-edge repair (TEER), offering lower rates of hospitalization for heart failure and overall mortality within 24 months of follow-up compared to standard medical therapy alone[1]. Given that a significant number of SMR patients exhibit LVEF<50% or present elevated surgical risks, TEER has emerged as a cornerstone in valvular heart failure treatment for this susceptible population. The preference for TEER is largely influenced by the positive outcomes seen in the COAPT trial and the less-than-favorable results of surgical MV repair for this specific concern. Therefore, the key point for decision-making in this group of patients is to select those patients who will benefit the most from TEER. Increasing researches underscore the importance of careful patient selection prior to TEER to further enhance outcomes[2-5]. However, the predictors of clinical improvement following TEER, remain ambiguous.

Echocardiographic assessment is critical for patients with TEER. Recent evidence suggests that right ventricular (RV) function serves as a critical predictor of heart failure progression[5]. Patients with RV pressure/volume overload or biventricular failure have a significantly increased 2-year mortality risk and more severe heart failure symptoms at follow-up. The newer echocardiographic metric, myocardial work extracted from pressure-strain loops (PSL), considers the dynamic interplay between internal heart pressures and myocardial contractions, offering supplementary data to strain, which is influenced by load conditions. And specifically, the evaluation of RV function using PSL provides important insights into the patients’ likely response to treatment[6, 7]. In this context, we present an investigation aimed to further understand the association between the changes in right ventricular myocardial work (RVMW) parameters following TEER and clinical improvement in HFrEF patients.

The aims of our study were: Firstly, to analyze the variations in RVMW parameters derived by PSL in HFrEF patients undergoing TEER; and secondly, to assess the association between immediate changes in these parameters and clinical improvement in patients undergoing TEER for SMR. We focused on patients who remained symptomatic despite optimal doses of guideline-directed medical therapy and cardiac resynchronization therapy (if appropriate), and who were deemed suitable for TEER by their cardiologist and unfit for mitral valve surgery by the cardiothoracic surgeon.

This research is critical as it seeks to provide insights into the potential value of RVMW as an evaluative measure to determine the effectiveness of TEER. Through this study, we aim to contribute to the evolving body of knowledge aimed at enhancing therapeutic strategies and patient outcomes in the complex field of heart failure management.

## Methods

### Study population

This prospective, observational, single-center study was conducted at our hospital from May 2021 to January 2023. The study was designed to monitor the change in RVMW parameters following TEER treatment in HFrEF patients and assess their association with clinical improvement. The study protocol was approved by the institutional research and ethical committees of Sichuan Provincial People’s Hospital and complied with the Declaration of Helsinki. All patients gave written informed consent before participation.

Eligible patients were adults with ischemic or nonischemic cardiomyopathy and left ventricular ejection fraction (LVEF) ranging from 20% to 50%. Participants were required to have moderate-to-severe (grade 3+) or severe (grade 4+) secondary mitral regurgitation (SMR) that had been confirmed by echocardiography. Patients were included if they remained symptomatic (NYHA functional class Ⅱ, Ⅲ, or Ⅳa [ambulatory]) despite optimal doses of guideline-directed medical therapy and cardiac resynchronization therapy (if appropriate), as per professional societies’ guidelines. Patients were excluded if they were deemed anatomically ineligible for device implantation by the interventional cardiologist or if the cardiothoracic surgeon deemed mitral valve surgery appropriate. Selected participants underwent the TEER procedure (MitraClipⓇ, Abbott structural, Santa Clara, CA, USA), which was performed by a team of interventional cardiologists.

### Echocardiography assessments

Utilizing the Vivid E95 ultrasound systems (General Electric Healthcare, Horten, Norway) equipped with M5S and 4Vc probes, comprehensive 2D echocardiography was conducted both prior to and immediately following the TEER procedure. For strain analyses, cine-loops spanning 40-90 frames per second over three consistent cycles during breath holds were recorded, focusing on the RV apical four-chamber views. This allowed for the automated tracking of acoustic marker positional changes. To calculate RV volumes, 3D full-volume cine-loops were secured, adjusted to encompass the RV based on a 12 multi-slice guide. To eliminate stitching artifacts, careful optimization was done to achieve a 1-beat acquisition at 20 volumes per second. The 3D dataset’s image quality was visually assessed, taking into account the signal-to-noise ratio and the existence of any dropouts or artefacts.

Standard parameters like tricuspid annular peak systolic excursion (TAPSE) via M-Mode, tricuspid annular systolic velocity (S’) through tissue Doppler, and RV fractional area change (FAC) – computed as [(RV end-diastolic area - RV end-systolic area) / RV end-diastolic area] × 100 – were executed as per established guidelines[8]. Custom software (EchoPAC version 204, General Electric, Horten Norway) was employed to determine speckle tracking data. Automated function imaging, with potential manual adjustments to the region of interest, was applied to ensure wall thickness was accurately captured for determining RV longitudinal strain (LS). The RV myocardial work index (MWI) was derived using specialized software originally crafted for LV myocardial work assessment by Russell and colleagues[9]. As previously outlined, the RV myocardial force-segment length loop’s area was equated with systolic pulmonary artery pressure (PAPs) (acting as a surrogate for myocardial force) - RV strain (acting as a surrogate for segment length) loops[6]. PAPs was directly gauged during right heart catheterization, while strain was ascertained as the mean LS of the RV free-wall and the interventricular septum. To craft a comprehensive bull’s eye map via automated function imaging, RV LS data derived from the RV-focused apical four-chamber view was repeated thrice as earlier reported[6, 7]. Given that the left ventricle contributes approximately 20-40% of the total RV stroke volume and pulmonary flow through the septum, the analysis centered on RV global instead of free wall strain. By aligning measurements of RV strain with pulmonary systolic and diastolic pressures based on cardiac cycle timings (gauged by pulmonic and tricuspid valve actions), non-invasively derived PSLs for the right ventricle were produced[6]. Four distinct RVMW parameters were extracted: (1) RV global work index (RVGWI, mmHg%), which is the area within the global RV pressure-strain loop, measured from tricuspid valve closure to its opening; (2) RV global constructive work (GCW, mmHg%), signifying the work aiding in cardiac myocyte systolic shortening and isovolumic relaxation lengthening; (3) RV global wasted work (GWW, mmHg%), representing the work resulting in cardiac myocyte systolic lengthening and isovolumic relaxation shortening; (4) RV global work efficiency (GWE, %), calculated as RVGCW divided by the combined total of RVGCW and RVGWW. To observe alterations in right ventricular myocardial work (RVMW) parameters, participants underwent echocardiographic assessments both before and right after the TEER procedure.

### Clinical improvement evaluation

Followed up was conducted at 30 days, 6 months, and 1 year. Clinical improvement was evaluated through the Kansas City Cardiomyopathy Questionnaire Overall Summary Score (KCCQ-OS) and the 6-min walk distance (6MWD) test at baseline and at 1-year after TEER. The KCCQ-OS assesses five domains of health status, including physical limitation, symptoms, quality of life, social limitation, and self-efficacy[10]. Scores for the KCCQ-OS range from 0 to 100, with higher scores indicating better health status. The 6MWD test measures the distance a patient can walk on a flat surface, back and forth down a 30-m hallway, in a period of 6 min. It was administered using standardized methodology with patients walking at their own pace with rest stops as needed[11].

### Statistical Analysis

Continuous variables are presented as mean ± standard deviation (SD) or median (interquartile range [IQR]), as appropriate, while categorical variables are expressed as frequencies and percentages. Comparisons between pre- and post-TEER parameters were performed using the paired t-test or Wilcoxon signed-rank test, as appropriate. Associations between the immediate change in RVMW parameters and clinical improvement were examined using multivariable linear regression analysis, adjusting for potential confounders. Kaplan-Meier estimates were used to analyze time-to-event variables, and the exponential Greenwood method was used to calculate SE. A *P*-value < 0.05 was considered statistically significant. Statistical analyses were performed using SPSS version 25.0 (SPSS Inc., IBM Corp).

## Results

### Population characteristics

Among 55 HFrEF patients with SMR who underwent TEER procedure, 3 patients were excluded because of poor echocardiographic imaging window and 4 because of a lack of invasive PAPs data. And finally, our study enrolled 48 patients with HFrEF (median age 68, range 57-84 years) (Figure 1). The study population was at high surgical risk, with STS risk scores averaging 7.42 (4.24-9.5). Most patients presented with severe heart failure symptoms (NYHA functional class ≥ III in 42 patients or 87.5%). 34 patients (70.8%) were implanted with 1 clip, and 14 patients (29.2%) were implanted with 2 clips. Single leaflet device detachment (SLDA) happened in one patient. One case with transient bradycardia and inferior wall lead ST segment elevation happened because of air embolism. Patients’ characteristics are presented in table 1.

**Figure 1.**
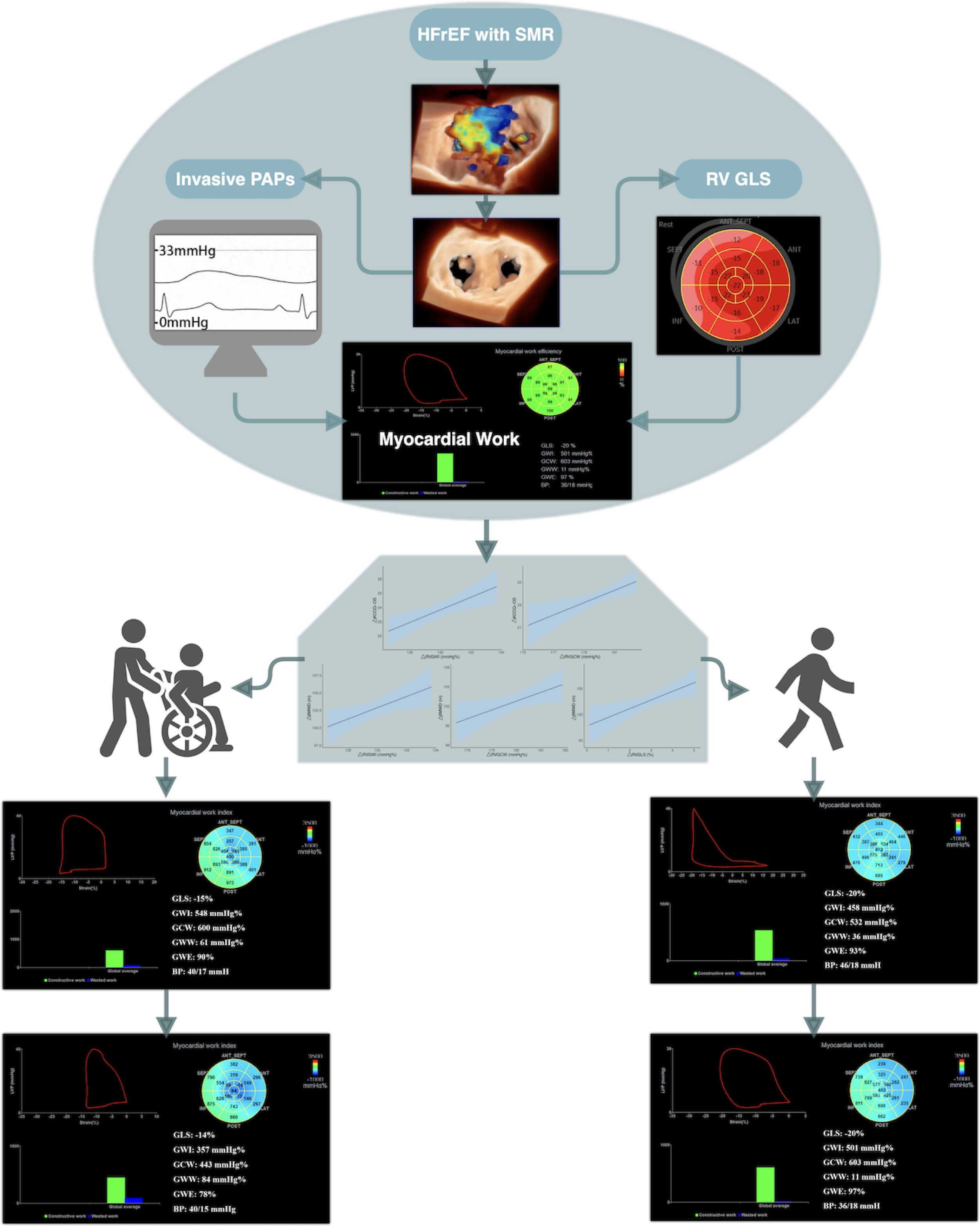
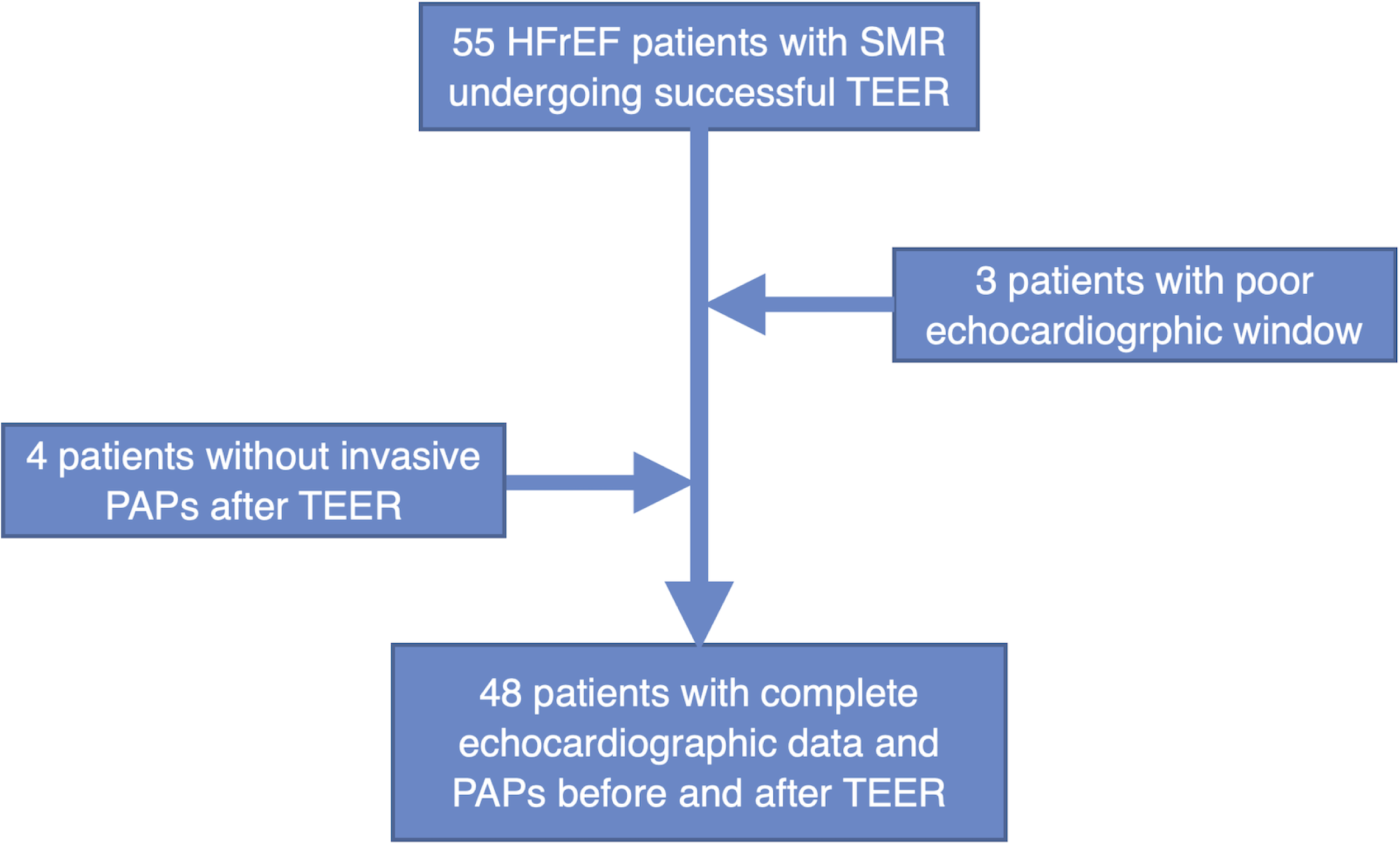
Study flow chart. HFrEF, heart failure with reduced ejection fraction; SMR, secondary mitral regurgitation; TEER, transcatheter edge-to-edge repair; PAPs, systolic pulmonary artery pressure

**Table 1.** patients characteristics (N=48) KCCQ-OS, Kansas City Cardiomyopathy Questionnaire Overall Summary Score; MPG, mean pressure gradient; STS, The Society of Thoracic Surgeons. Patients with coronary artery disease includes those with myocardial infarction, percutaneous coronary intervention, coronary artery bypass grafting, and any coronary artery stenosis>50% confirmed by coronary angiography or CTA

|  |  |
| --- | --- |
| Age, yrs | 68±15 |
| Male, % | 29, 60.4% |
| Body mass index, kg/m<sup>2</sup> | 24.7±2.7 |
| SBP, mmHg | 110±17 |
| DBP, mmHg | 63±10 |
| HR, bpm | 75±18 |
| eGFR, ml/min | 48.9±19.5 |
| NT-proBNP, pg/ml | 3325 (1845-5871) |
| STS risk score for MV repair | 7.42 (4.24-9.5) |
| Ischemic cardiomyopathy, % | 16, 33.3% |
| Coronary artery disease, % | 39, 81.3% |
| Prior myocardial infarction, % | 13, 27.1% |
| Prior coronary artery bypass grafting, % | 2, 4.2% |
| Peripheral vascular disease, % | 6, 12.5% |
| Diabetes, % | 20, 41.7% |
| Hypertension, % | 21, 43.8% |
| Atrial fibrillation or flutter | 20, 41.7% |
| NYHA class baseline |  |
| I | 0 |
| II | 6 (12.5%) |
| III | 20 (41.7%) |
| IV | 22 (45.8%) |
| NYHA class at follow-up |  |
| I | 6 (12.5%) |
| II | 29 (60.4%) |
| III | 10 (20.8%) |
| IV | 3 (6.3%) |
| Medication |  |
| RAS-I/ARNI | 31 (64.5) |
| Beta-blocker | 36 (75%) |
| MRA | 25 (52.1%) |
| Diuretic (ang type) | 41 (85.4%) |
| SGLT2i | 25 (52.1%) |
| Echocardiography |  |
| MR vena contracta, mm | 6.2±2.5 |
| MR EROA, cm<sup>2</sup> | 0.47±0.19 |
| MR RegVol, ml | 63.8±20.7 |
| MR severity baseline |  |
| 3+ | 19 (39.6%) |
| 4+ | 29 (60.4%) |
| AP systolic diameter, cm | 2.97±0.59 |
| AP diastolic diameter, cm | 3.48±0.62 |
| AL-PM systolic diameter, cm | 3.15±0.67 |
| AL-PM diastolic diameter, cm | 3.39±0.62 |
| LVEDV, ml | 187.5±57.1 |
| LVESV, ml | 128.6±50.1 |
| LVEDD, mm | 62.6±10.8 |
| LVESD, mm | 47.1±15.1 |
| LVEF, % | 32.7±6.9 |
| LAVI, ml/m<sup>2</sup> | 56.1±24.6 |
| TAPSE, mm | 16.5±5.1 |
| RV EDA, cm<sup>2</sup> | 23.9±6.8 |
| RV ESA, cm<sup>2</sup> | 14.8±5.2 |
| RV FAC, % | 37±14 |
| TR, m/s | 3.1±1.2 |
| SPAP-non-invasive, mmHg | 46.5±12.9 |
| SPAP-invasive before TEER, mmHg | 45.8±14.6 |
| SPAP-invasive after TEER, mmHg | 36.9±13.1 |
| TR severity baseline |  |
| 0+ | 0 |
| 1+ | 2 (4.2%) |
| 2+ | 10 (20.8%) |
| 3+ | 28 (58.3%) |
| 4+ | 8 (16.7%) |
| Mean gradient of MV before TEER (mmHg) | 1.8±0.7 |
| Mean gradient of MV after TEER (mmHg) | 3.8±1.5 |
| MPG ≤ 5mmHg | 45 (93.7%) |
| MPG ≥ 5mmHg | 3 (6.3%) |
| Coaptation depth (cm) | 0.87±0.31 |
| Coaptation depth ≤ 1.1cm | 39 (81.3%) |
| Coaptation depth ≥ 1.1cm | 9 (18.7%) |
| Coaptation length (cm) | 0.31±0.13 |
| Coaptation length ≤ 0.2cm | 8 (16.7%) |
| Coaptation length ≥ 0.2cm | 40 (83.3%) |
| Number of clips implanted |  |
| 1 clip | 34 (70.8%) |
| 2 clips | 14 (29.2%) |
| Type of clips |  |
| NTR | 27 |
| XTR | 26 |
| MR severity postimplant |  |
| 1+ | 25 (52.1%) |
| 2+ | 20 (41.7%) |
| ≥3+ | 3 (6.3%) |
| Baseline KCCQ-OS | 43.7±20.9 |
| KCCQ-OS at 1-year after TEER | 68.5±29.1 |
| Baseline 6MWD, m | 230 (185-323) |
| 6MWD at 1-year after TEER, m | 325 (260-380) |

### Echocardiographic characteristics

Following TEER procedure, MR severity was reduced to ≤ 2+ in 93.8% (45 patients) and ≤ 1+ in 52.1% (25 patients) (Figure 2). Mitral valve mean gradient increased from 1.8±0.7 mmHg at baseline to 3.8±1.5 mmHg post-procedure. Other procedural characteristics are presented in table 1. LVEDV decreased significantly post-procedure (from 187.5 ± 57.1 ml to 166 ± 48 ml, *p* < 0.001), with no significant change in LVESV (*p* > 0.05). LVEF also showed a decrease post-TEER (from 32.7 ± 6.9% to 29.8 ± 3.6%, *p* = 0.043). LVGLS also decreased after procedure (from -12.9±3.8% to -10.2±2.1%, *p* = 0.036). LAVI decreased from 56.1±24.6 ml/m^2^ before to 53.2±11.5 ml/m^2^ after TEER (*p* = 0.070). LVGWI, LVGCW increased (658±314mmHg% and 871±356mmHg% vs 738±281mmHg% and 917±349mmHg%, *p* = 0.028 and 0.039, respectively), while no significant change was observed in LVGWW and LVGWE after TEER (139±37mmHg% and 85.5±8.1% vs 151±43mmHg% and 84.4±6.7%, *p* = 0.059 and 0.225, respectively). (Table 2).

**Figure 2.**
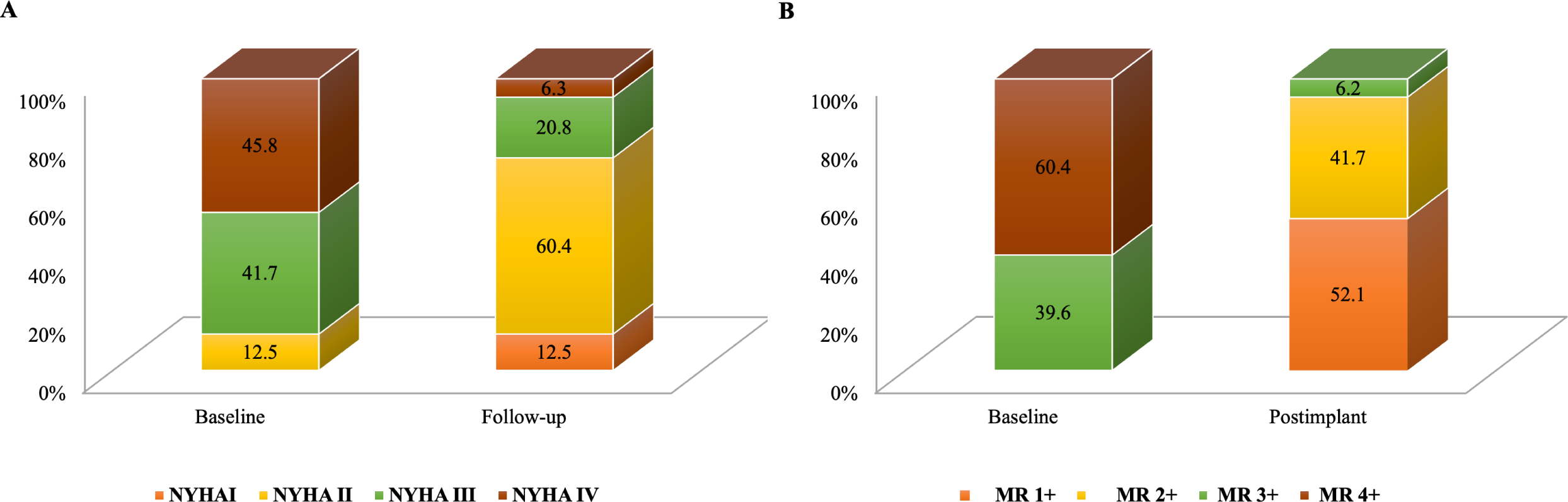
Distributions of mitral regurgitation severity (A) and New York Heart Association functional class (B) at baseline and follow-up.

**Table 2.** Echocardiographic parameters at baseline and immediately after TEER.

|  | Baseline | immediately after<br>TEER | Parameter<br>changes | *P* |
| --- | --- | --- | --- | --- |
| LVEDV (ml) | 187.5±57.1 | 166 ± 48 | -20.5±6.9 | <0.001 |
| LVESV (ml) | 128.6±50.1 | 121 ± 47 | -6.8±2.1 | 0.065 |
| LVEF (%) | 32.7±6.9 | 29.8±3.6 | -2.1±0.3 | 0.043 |
| LVGLS (%) | -12.9±3.8 | -10.2±2.1 | 2.3±1.2 | 0.036 |
| LVGWI<br>(mmHg%) | 658±314 | 738±281 | 75±23 | 0.028 |
| LVGCW<br>(mmHg%) | 871±356 | 917±349 | 49±21 | 0.039 |
| LVGWW<br>(mmHg%) | 139±37 | 151±43 | 10±4 | 0.059 |
| LVGWE (%) | 85.5±8.1 | 84.4±6.7 | -1.3±0.4 | 0.225 |
| LAVI (ml/m<sup>2</sup>) | 56.1±24.6 | 53.2±11.5 | -3.5±0.7 | 0.070 |
| RVEDV (ml) | 128.5 ± 20.7 | 136.7 ± 28.5 | 8.1±3.6 | 0.063 |
| RVESV (ml) | 82.9 ± 17.6 | 73.5 ± 20.6 | -10±4 | 0.065 |
| RVSV (ml) | 44.5 ± 9.8 | 58.8 ± 14.5 | 15.8±6.8 | 0.023 |
| RVEF (%) | 36.6±3.6 | 45.4±3.9 | 8.7±3.2 | 0.008 |
| RVFAC (%) | 37±14 | 39±12 | 2.5±1.4 | 0.120 |
| TAPSE (mm) | 16.5±5.1 | 17.1±2.1 | 0.7±0.3 | 0.130 |
| RV GLS (%) | -9.7±3.8 | -12.5±3.5 | -3.0±0.4 | 0.025 |
| RV S' (m/s) | 6.7±2.2 | 6.9±1.8 | 0.17±0.06 | 0.350 |
| RVGWI<br>(mmHg%) | 452.4±112.5 | 589.4±119.6 | 135.1±29.8 | 0.030 |
| RVGCW<br>(mmHg%) | 596.3±127.5 | 778.8±135.3 | 180.5±32.1 | 0.025 |
| RVGWW<br>(mmHg%) | 62±19.7 | 63±18.7 | 1.4±0.5 | 1.410 |
| RVGWE (%) | 85.7±15.6 | 91.2±22.4 | 6.2±2.3 | 0.037 |

There’s no significant change in RVEDV (128.5 ± 20.7ml vs 136.7 ± 28.5ml, *p* = 0.063), RVESV (82.9 ± 17.6ml vs 73.5 ± 20.6ml, *p* = 0.065) after TEER. While RVSV (44.5 ± 9.8ml vs 58.8 ± 14.5ml, *p* = 0.023) and RVEF (36.6±3.6% vs 45.4±3.9%, *p* = 0.008) increased. TAPSE, RV S’and RV FAC were not significantly changed immediately after MitraClip treatment (16.5±5.1 mm, 6.7±2.2m/s and 37±14% before vs 17.1±2.1 mm, 6.9±1.8m/s and 39±12% after MitraClip treatment, all *p* > 0.05). While, RV GLS, RVGWI, RVGCW, and RVGWE were significantly increased after MitraClip treatment (-9.7±3.8%, 452.4±112.5 mmHg%, 596.3±127.5 mmHg%, and 85.7±15.6% before vs -12.5±3.5%, 589.4±119.6 mmHg%, 778.8±135.3 mmHg%, and 91.2±22.4% after MitraClip treatment, *p* = 0.025, 0.030, 0.025 and 0.037, respectively). In contrast, no significant change was observed in RVGWW after MitraClip treatment (62±19.7 mmHg% vs 63±18.7 mmHg%, *p* > 0.05). Other left and right ventricular morphologic and functional parameters are presented in table 2.

### Clinical outcomes at follow-up

During follow-up, 2 patients (4.2%) died because of non-cardiovascular disease. The total number of hospitalizations for heart failure during follow up was 5 (10.4%), in which one patient underwent left ventricular assist device (LVAD) implantation. And there are no other major adverse cardiovascular events during follow-up. NYHA functional class at follow-up (12-month) was ≤ Ⅱ in 72.9% (35 patients) (Figure 2). NT-proBNP decreased significantly from 3325 (1845-5871) pg/ml at baseline to 1025 (450-1870) pg/ml at 1 year. Patient health status, as measured by KCCQ-OS and 6MWD, improved at 30 days, and this improvement continued at 12-month (mean KCCQ-OS increased from 43.7±20.9 to 68.5±29.1 points at 12-month after implantation, and mean 6MWD from 230 (185-323) m to 325 (260-380) m at 12-month after implantation) (Figure 3). The Kaplan-Meier estimates for survival, freedom from HF hospitalization at 12 months were 95.8% and 89.1%, respectively (Figure 4).

**Figure 3.**
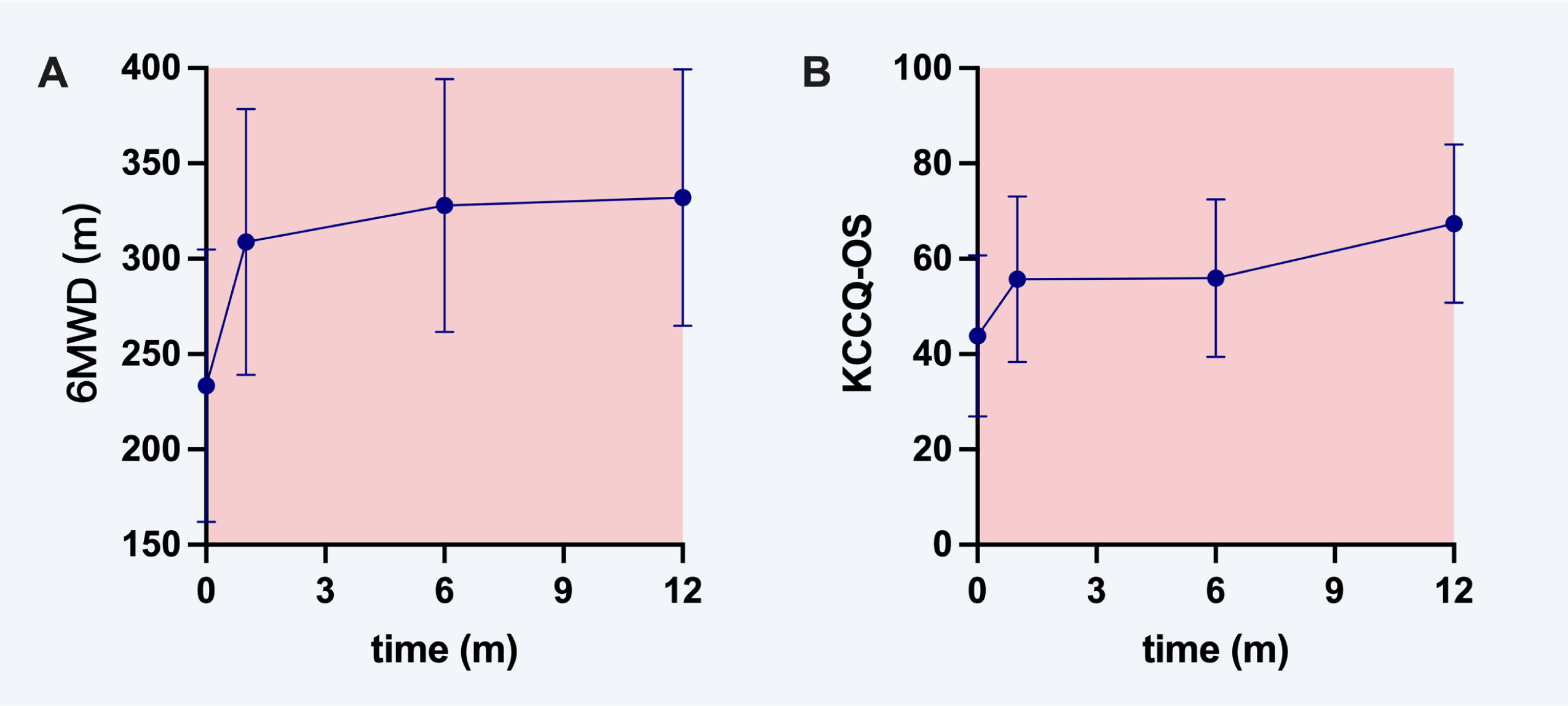
6WMD (A) and KCCQ-OS (B) changes from baseline to 1-year follow-up after TEER. KCCQ-OS, Kansas City Cardiomyopathy Questionnaire Overall Summary Score; 6MWD, 6-minute walk distance test.

**Figure 4.**
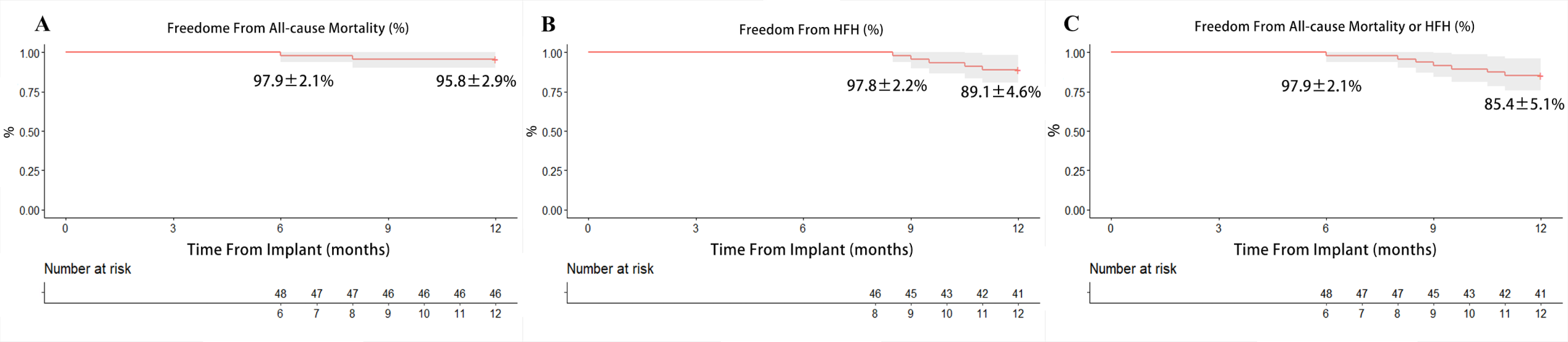
Kaplan-Meier estimates for freedom from (A) all-cause mortality, (B) heart failure hospitalization (HFH), and (C) all-cause mortality and HFH. Graphs show Kaplan-Meier estimate ± SE and error bars represent 95% CI.

On multivariable linear regression analysis, RVGWI and RVGCW immediate change was independently associated with KCCQ-OS (△RVGWI: ***β*** = 0.40, *P* < 0.001; △RVGCW: ***β*** = 0.39, *P* =0.003), RVGWI, RVGCW and RVGLS immediate change were independently associated with 6MWD improvement (△RVGWI: ***β*** = 0.31, *P* = 0.029; △RVGCW: ***β*** = 0.30, *P* = 0.039; △RVGLS: ***β*** = 0.35, *P* = 0.041) (Table 3, Figure 5 and 6).

**Figure 5.**
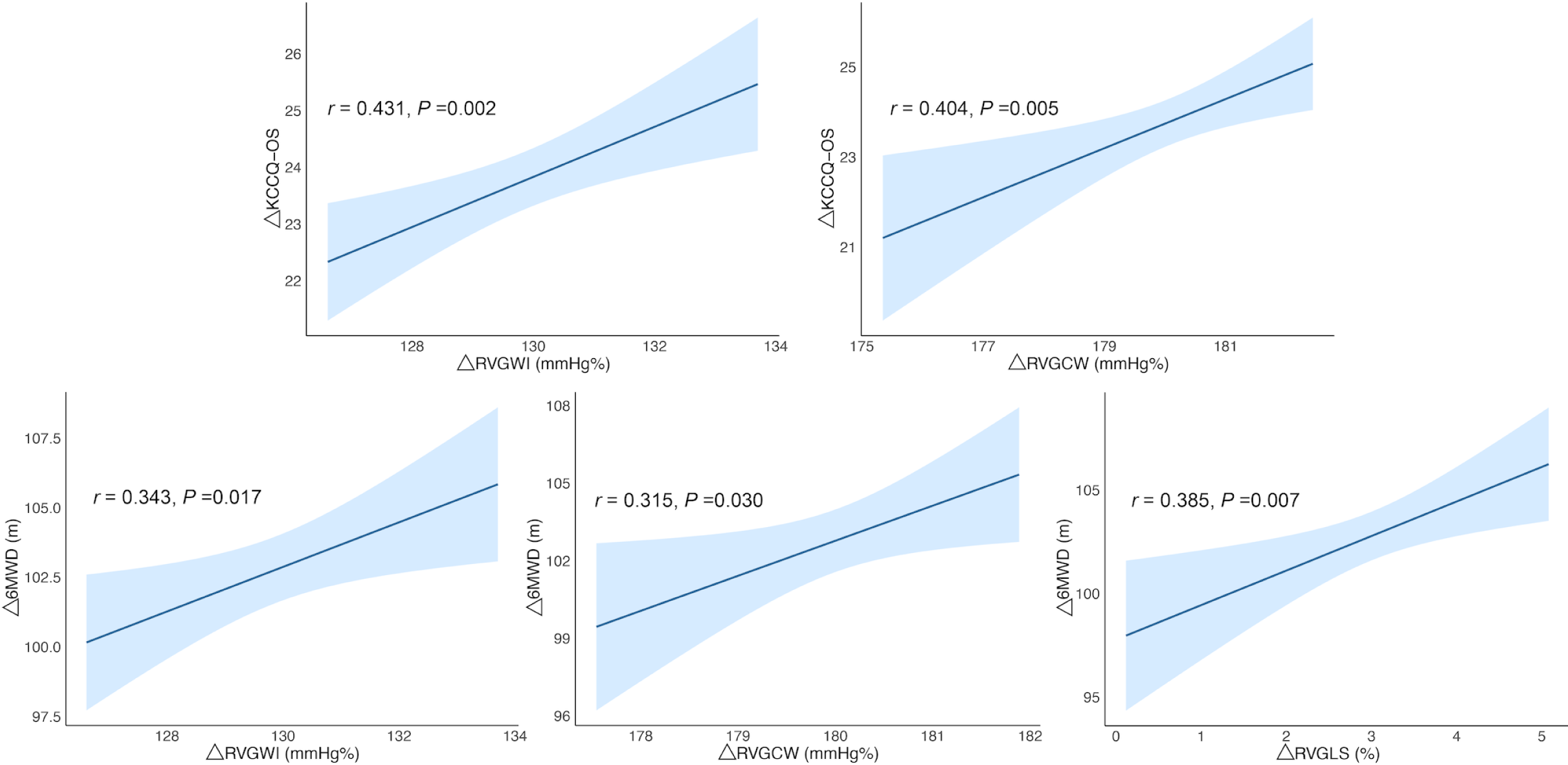
Correlation between clinical improvement and RVGWI, RVGCW, RVGLS changes. KCCQ-OS, Kansas City Cardiomyopathy Questionnaire Overall Summary Score; 6MWD, 6-minute walk distance test; RVGWI, right ventricular global work index; RVGCW, right ventricular global constructive work; RVGLS, right ventricular global longitudinal strain.

**Figure 6.**
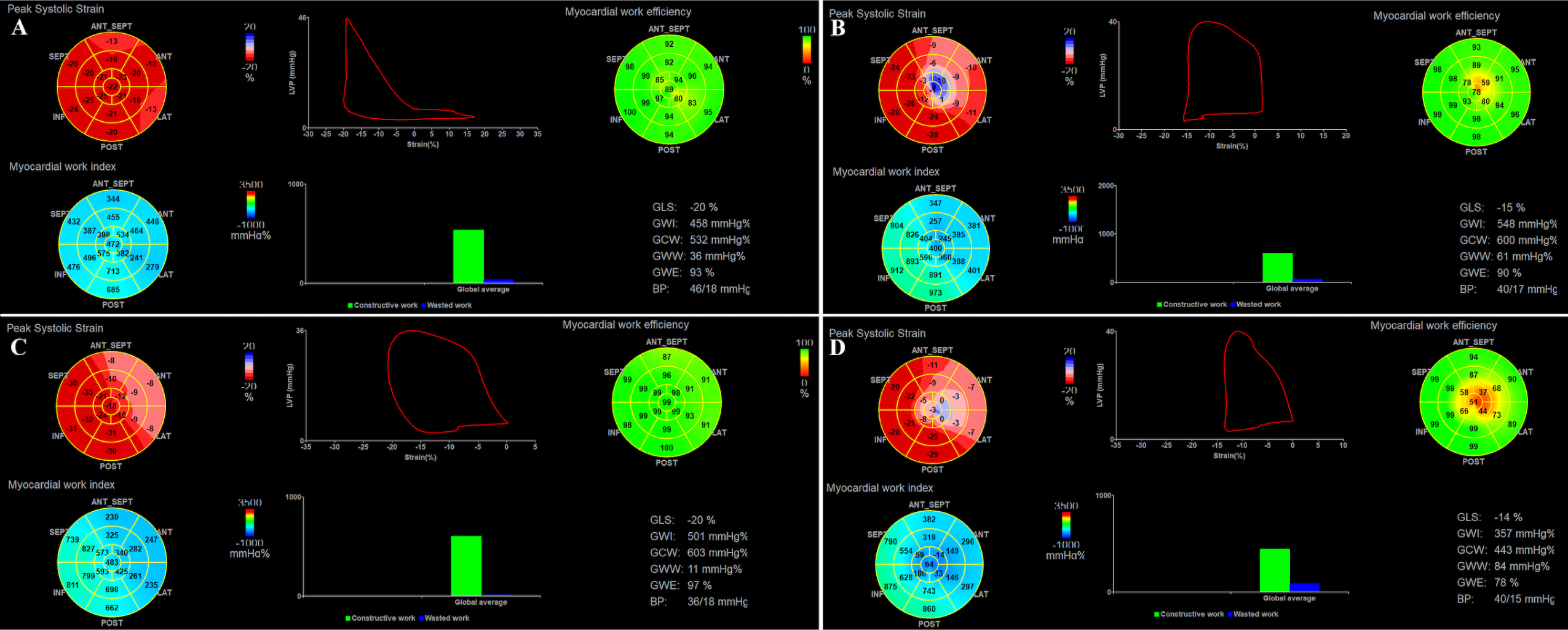
Two patients with and without clinical improvement after TEER. One patient with obvious clinical improvement after TEER showed significant increase in RVGWI and RVGCW (A, C). While RVGWI and RVGCW decreased after TEER in another patient without significant clinical improvement after TEER (B, D).

**Table 3.** predictor of KCCQ-OS and 6MWD improvement at follow-up.

|  | KCCQ-OS |  | 6MWD |  |
| --- | --- | --- | --- | --- |
|  | Univariable | Multivariable | Univariable | Multivariable |

|  | analysis |  | analysis |  | analysis |  | analysis |  |
| --- | --- | --- | --- | --- | --- | --- | --- | --- |
| | *r* | *P* | $\beta$ | *P* | *r* | *P* | $\beta$ | *P* |
| ΔLVEDV<br>(ml) | -0.231 | 0.070 | | | -0.332 | 0.041 | | |
| ΔLVESV<br>(ml) | -0.072 | 0.569 | | | -0.052 | 0.673 | | |
| ΔLVEF (%) | -0.213 | 0.156 | | | -0.123 | 0.594 | | |
| ΔLVGLS<br>(%) | 0.353 | 0.044 | | | 0.494 | 0.035 | | |
| ΔLVGWI<br>(mmHg%) | 0.373 | 0.057 | | | 0.291 | 0.096 | | |
| ΔLVGCW<br>(mmHg%) | 0.294 | 0.04 | | | 0.385 | 0.032 | | |
| ΔLVGWW<br>(mmHg%) | -0.075 | 0.733 | | | -0.093 | 0.673 | | |
| ΔLVGWE<br>(%) | -0.166 | 0.256 | | | -0.246 | 0.134 | | |
| ΔLAVI<br>(ml/m<sup>2</sup>) | 0.237 | 0.077 | | | 0.257 | 0.096 | | |
| ΔRVEDV<br>(ml) | -0.198 | 0.242 | | | -0.105 | 0.293 | | |
| ΔRVESV<br>(ml) | 0.319 | 0.064 | | | 0.357 | 0.036 | | |
| ΔRVSV<br>(ml) | 0.297 | 0.045 | | | 0.184 | 0.083 | | |
| ΔRVEF (%) | 0.188 | 0.226 | | | 0.257 | 0.154 | | |
| ΔRVFAC<br>(%) | 0.375 | 0.087 | | | 0.352 | 0.043 | | |
| ΔTAPSE<br>(mm) | 0.284 | 0.034 | | | 0.294 | 0.024 | | |
| ΔRV GLS<br>(%) | 0.255 | 0.024 | | | 0.385 | 0.007 | 0.350 | 0.041 |
| ΔRV | 0.392 | 0.032 | | | 0.376 | 0.025 | | |
| S'(m/s) |  |  |  |  |  |  |  |  |
| ΔRVGWI<br>(mmHg%) | 0.431 | 0.002 | 0.397 | <0.001 | 0.343 | 0.017 | 0.310 | 0.029 |
| ΔRVGCW<br>(mmHg%) | 0.404 | 0.005 | 0.390 | 0.003 | 0.315 | 0.030 | 0.300 | 0.039 |
| ΔRVGWW<br>(mmHg%) | -0.473 | 0.047 | | | -0.104 | 0.090 | | |
| ΔRVGWE<br>(%) | 0.413 | 0.057 | | | 0.394 | 0.043 | | |

## Discussion

Our research illuminates the assessment of RV function in TEER patients, introducing several groundbreaking insights. This is the first studies to use PSL to quantify and compare immediate change of RVMW before and after TEER. This is also the first research to study the association of RVMW changes after TEER and clinical improvement. The main findings including: (1) Traditional RV systolic function parameters were not significantly changed immediately after MitraClip treatment, such as TAPSE, RV GLS, RV S’ and RV FAC RVMW. (2) RVGWI, RVGCW, and RVGWE were significantly increased at post procedure. (3) RVMW was an important predictor of clinical improvement in HFrEF patients undergoing TEER.

### Superiority of RVMW over standard RV systolic function parameters

Differing from RV longitudinal strain, TAPSE, and RV FAC, RVMW parameters amalgamate aspects like contractility, RV dissynchrony, and pulmonary pressures. Beyond offering a holistic evaluation of RV function, RVMW isn’t constrained by the technical limitations inherent to other standard RV systolic function metrics. TAPSE, for instance, is influenced by angles, load, and the extent of cardiac translation, whereas RV FAC’s accuracy is hampered by its load-dependency and only moderate interobserver reproducibility[8, 12]. While both experimental and clinical investigations have highlighted that RV longitudinal strain, gauged via speckle tracking echocardiography, is affected by afterload, it’s arguably less influenced than other conventional RV systolic function measures[12]. In our study, RVEF and RVGLS significantly increased after TEER, while other conventional RV functional parameters didn’t change obviously.

The RV-to-pulmonary artery (RV-PA) coupling, non-invasively gauged via echocardiography by analyzing the ratio between tricuspid annular plane systolic excursion and pulmonary artery systolic pressure (TAPSE/PASP), can be viewed as an approximate for the RV length-force relationship and has been found to hold clinical and prognostic significance in HF demographics[13-16]. Adamo and his colleagues found that TAPSE/PAPs is a major predictor of outcome in patients with SMR undergoing TEER[17]. Given its consideration for afterload, RVMW offers insights into RV-PA coupling, potentially offering a more nuanced understanding of RV systolic function. From RV-PA coupling point of view, our result complies with previous study, which showed improvement in TAPSE/PAPs after successful TEER is associated with a better outcome[17]. Initial evidence supporting the application of RVMW to the RV was documented using PAPs deduced from TR velocity[6]. In our approach, we opted for invasive PAPs measurements to ensure precision.

### Changes in RV functional parameters

Given a compromised left ventricle, pulmonary hypertension and RV dysfunction are closely intertwined. SMR amplifies pulmonary pressures, thus exacerbating RV failure. Patients with HFrEF coupled with pronounced SMR tend to exhibit more pronounced RV remodeling compared to those with milder SMR[18]. Alleviating a component of RV overload via SMR correction can potentially bolster RV contractility. Multiple concise studies have pinpointed immediate benefits post TEER in terms of pulmonary hypertension regression and RV dysfunction recovery, with the enhancements linked to superior outcomes[19-24]. However, literature on the trajectory of RVMW post TEER is scarce, and juxtapositions of RVMW alterations with other traditional RV function parameter shifts after TEER remain uncharted. As far as our understanding goes, our study is the first to probe into the shifts in RVMW metrics post TEER. Our findings spotlight a significant surge in RVMW parameters, encompassing RVGWI, RVGCW, and RVGWE, post TEER, likely attributed to reduced pulmonary congestion given the presence of RV contractile reserve.

### Association of changes in RVMW and clinical improvement

The COAPT trial demonstrated the benefit of TEER in reducing hospitalizations for heart failure and mortality in HFrEF patients with moderate-to-severe or severe SMR, which is corroborated by our findings[25]. However, the selection of patients who would derive maximum benefit from TEER remains a challenge. Our study provides significant insights into this aspect by evaluating the association between immediate change of right ventricular function and clinical improvement post-TEER.

In line with prior studies, right ventricular function emerged as a crucial predictor for heart failure, underscoring the importance of comprehensive assessment of both ventricles in managing HFrEF patients. While conventional parameters such as TAPSE, FAC, and RV GLS did not show significant change post-TEER, RVMW parameters effectively captured the functional improvement.

In our study, the immediate change in these parameters was independently associated with the clinical improvement evaluated through KCCQ-OS and 6MWD. These findings suggest that improvements in RVMW parameters could potentially serve as predictive markers for clinical improvement following TEER. The non-invasive RVMW analysis we proposed proved to be easy to perform and could provide essential insights about the RV reserve function and its impact on patient outcomes post-TEER. This is particularly relevant considering the challenging nature of heart failure management, where RV function often plays a critical role.

Despite the reduction in LVEF post-TEER, we observed a marked increase of LVMW, RVMW parameters and clinical improvement, reflecting the complexity of heart failure pathophysiology. And only RVMW changes were independently associated with clinical improvement. This suggests that RVMW parameters might provide a more holistic assessment of cardiac function in HFrEF patients with TEER than LV functional measures and other traditional RV functional parameters.

### Study limitations

Our study does come with its set of constraints. Firstly, the sample size we worked with was on the smaller side, potentially affecting the wider applicability of our findings. Additionally, the proprietary software utilized for RVMW measurements is exclusive to a single vendor and was originally crafted for analyzing the left ventricle myocardial work. The creation of LV pressure-strain loops relies on Laplace’s law, which operates on basic geometric presumptions. Given the intricate and irregular geometry of the RV, the myocardial work calculations derived might not be as accurate as those for the LV[26]. It might be prudent to corroborate non-invasive RV PSL with pressure-volume loops acquired invasively. While we did account for a range of potential confounding variables, we cannot completely dismiss the possibility of unobserved confounders influencing the results. To bolster the robustness of our conclusions and to gain insights into the long-term ramifications of RVMW changes post-TEER, future research endeavors would benefit from a more expansive participant pool and extended observation durations.

## Conclusion

Our investigation revealed a significant increase in RVMW after the application of a MitraClip treatment in HFrEF patients who underwent this treatment for SMR. More importantly, we found that an increase in RVGWI and RVGCW was independently associated with clinical improvement in these patients. This suggests that RV reserve function could be vital predictors for assessing the clinical outcome of TEER in HFrEF patients with SMR. Furthermore, our findings hint at a potential optimization of the therapeutic strategy for HFrEF patients undergoing TEER, by taking into account RV function and myocardial work parameters. Future research is needed to further validate these findings in larger patient cohorts and to further refine the use of RVMW parameters in the clinical decision-making process for TEER.

## Data Availability

All data referred to this manuscript are available from the corresponding author (Yi Wang) upon reasonable request.

## Acknowledgement

The authors would like to thank our study nurse Pei Fu for her dedicated support at patient follow-up visits and data acquisition.

## Reference

1. Orban, M., W. Rottbauer, M. Williams, P. Mahoney, R.S. von Bardeleben, M.J. Price, et al. Transcatheter edge-to-edge repair for secondary mitral regurgitation with third-generation devices in heart failure patients - results from the Global EXPAND Post-Market study. Eur J Heart Fail 2023; 25(3): 411–421.

2. Doldi, P.M., J. Buech, M. Orban, P. Samson-Himmelstjerna, U. Wilbert-Lampen, C. Hagl, et al. Transcatheter mitral valve repair may increase eligibility for heart transplant listing in patients with end-stage heart failure and severe secondary mitral regurgitation. Int J Cardiol 2021; 338: 72–78.

3. Karam, N., L. Stolz, M. Orban, S. Deseive, F. Praz, D. Kalbacher, et al. Impact of Right Ventricular Dysfunction on Outcomes After Transcatheter Edge-to-Edge Repair for Secondary Mitral Regurgitation. JACC Cardiovasc Imaging 2021; 14(4): 768–778.

4. Koell, B., M. Orban, J. Weimann, M. Kassar, N. Karam, M. Neuss, et al. Outcomes Stratified by Adapted Inclusion Criteria After Mitral Edge-to-Edge Repair. J Am Coll Cardiol 2021; 78(24): 2408–2421.

5. Stolz, L., P.M. Doldi, M. Orban, N. Karam, T. Puscas, M.G. Wild, et al. Staging Heart Failure Patients With Secondary Mitral Regurgitation Undergoing Transcatheter Edge-to-Edge Repair. JACC Cardiovasc Interv 2023; 16(2): 140–151.

6. Butcher, S.C., F. Fortuni, J.M. Montero-Cabezas, R. Abou, M. El Mahdiui, P. van der Bijl, et al. Right ventricular myocardial work: proof-of-concept for non-invasive assessment of right ventricular function. Eur Heart J Cardiovasc Imaging 2021; 22(2): 142–152.

7. Sade, L.E., A. Colak, S.A. Duzgun, T. Hazirolan, A. Sezgin, E. Donal, et al. Approach to optimal assessment of right ventricular remodelling in heart transplant recipients: insights from myocardial work index, T1 mapping, and endomyocardial biopsy. Eur Heart J Cardiovasc Imaging 2023; 24(3): 354-363.

8. Lang, R.M., L.P. Badano, V. Mor-Avi, J. Afilalo, A. Armstrong, L. Ernande, et al. Recommendations for cardiac chamber quantification by echocardiography in adults: an update from the American Society of Echocardiography and the European Association of Cardiovascular Imaging. Eur Heart J Cardiovasc Imaging 2015; 16(3): 233–70.

9. Russell, K., M. Eriksen, L. Aaberge, N. Wilhelmsen, H. Skulstad, E.W. Remme, et al. A novel clinical method for quantification of regional left ventricular pressure-strain loop area: a non-invasive index of myocardial work. Eur Heart J 2012; 33(6): 724–33.

10. Green, C.P., C.B. Porter, D.R. Bresnahan, and J.A. Spertus. Development and evaluation of the Kansas City Cardiomyopathy Questionnaire: a new health status measure for heart failure. J Am Coll Cardiol 2000; 35(5): 1245–55.

11. Laboratories, A.T.S.C.o.P.S.f.C.P.F. ATS statement: guidelines for the six-minute walk test. Am J Respir Crit Care Med 2002; 166(1): 111–7.

12. Tadic, M., E. Pieske-Kraigher, C. Cuspidi, D.A. Morris, F. Burkhardt, A. Baudisch, et al. Right ventricular strain in heart failure: Clinical perspective. Arch Cardiovasc Dis 2017; 110(10): 562–571.

13. Guazzi, M., R. Naeije, R. Arena, U. Corra, S. Ghio, P. Forfia, et al. Echocardiography of Right Ventriculoarterial Coupling Combined With Cardiopulmonary Exercise Testing to Predict Outcome in Heart Failure. Chest 2015; 148(1): 226–234.

14. Ghio, S., M. Guazzi, A.B. Scardovi, C. Klersy, F. Clemenza, E. Carluccio, et al. Different correlates but similar prognostic implications for right ventricular dysfunction in heart failure patients with reduced or preserved ejection fraction. Eur J Heart Fail 2017; 19(7): 873–879.

15. Guazzi, M., D. Dixon, V. Labate, L. Beussink-Nelson, F. Bandera, M.J. Cuttica, et al. RV Contractile Function and its Coupling to Pulmonary Circulation in Heart Failure With Preserved Ejection Fraction: Stratification of Clinical Phenotypes and Outcomes. JACC Cardiovasc Imaging 2017; 10(10 Pt B): 1211-1221.

16. Santas, E., P. Palau, M. Guazzi, R. de la Espriella, G. Minana, J. Sanchis, et al. Usefulness of Right Ventricular to Pulmonary Circulation Coupling as an Indicator of Risk for Recurrent Admissions in Heart Failure With Preserved Ejection Fraction. Am J Cardiol 2019; 124(4): 567–572.

17. Adamo, M., R.M. Inciardi, D. Tomasoni, L. Dallapellegrina, R. Estevez-Loureiro, D. Stolfo, et al. Changes in Right Ventricular-to-Pulmonary Artery Coupling After Transcatheter Edge-to-Edge Repair in Secondary Mitral Regurgitation. JACC Cardiovasc Imaging 2022; 15(12): 2038–2047.

18. Goliasch, G., P.E. Bartko, N. Pavo, S. Neuhold, R. Wurm, J. Mascherbauer, et al. Refining the prognostic impact of functional mitral regurgitation in chronic heart failure. Eur Heart J 2018; 39(1): 39–46.

19. Giannini, C., A.S. Petronio, M. De Carlo, F. Guarracino, L. Conte, F. Fiorelli, et al. Integrated reverse left and right ventricular remodelling after MitraClip implantation in functional mitral regurgitation: an echocardiographic study. Eur Heart J Cardiovasc Imaging 2014; 15(1): 95–103.

20. Godino, C., A. Salerno, M. Cera, E. Agricola, G. Fragasso, I. Rosa, et al. Impact and evolution of right ventricular dysfunction after successful MitraClip implantation in patients with functional mitral regurgitation. Int J Cardiol Heart Vasc 2016; 11: 90–98.

21. Ledwoch, J., C. Fellner, P. Hoppmann, R. Thalmann, H. Kossmann, M. Dommasch, et al. Impact of transcatheter mitral valve repair using MitraClip on right ventricular remodeling. Int J Cardiovasc Imaging 2020; 36(5): 811–819.

22. Sauter, R.J., J. Patzelt, M. Mezger, H. Nording, J.C. Reil, M. Saad, et al. Conventional echocardiographic parameters or three-dimensional echocardiography to evaluate right ventricular function in percutaneous edge-to-edge mitral valve repair (PMVR). Int J Cardiol Heart Vasc 2019; 24: 100413.

23. Caiffa, T., A. De Luca, E. Biagini, L. Lupi, F. Bedogni, M. Castrichini, et al. Impact on clinical outcomes of right ventricular response to percutaneous correction of secondary mitral regurgitation. Eur J Heart Fail 2021; 23(10): 1765–1774.

24. Stolfo, D., A. De Luca, G. Morea, M. Merlo, G. Vitrella, T. Caiffa, et al. Predicting device failure after percutaneous repair of functional mitral regurgitation in advanced heart failure: Implications for patient selection. Int J Cardiol 2018; 257: 182–187.

25. Stone, G.W., J. Lindenfeld, W.T. Abraham, S. Kar, D.S. Lim, J.M. Mishell, et al. Transcatheter Mitral-Valve Repair in Patients with Heart Failure. N Engl J Med 2018; 379(24): 2307–2318.

26. Redington, A.N., H.H. Gray, M.E. Hodson, M.L. Rigby, and P.J. Oldershaw. Characterisation of the normal right ventricular pressure-volume relation by biplane angiography and simultaneous micromanometer pressure measurements. Br Heart J 1988; 59(1): 23–30.

